# Occupational determinants of COVID-19 cases and vaccination: an ecological analysis of counties in the United States as of December 2021

**DOI:** 10.1101/2023.01.13.22274536

**Authors:** John S. Ji, Yucheng Wang, Dustin T. Duncan

## Abstract

**Objective:** We aim to study the relationship between occupation distribution within each county and COVID-19 cumulative incidence and vaccination rate in the United States.

**Methods:** We collected county-level data from January 22, 2020 up to December 25, 2021. We fit multivariate linear models to find the relationship of the percentage of people employed by 23 main occupations.

**Results:** Counties with more health-related jobs, office support roles, community service, sales, production and material moving occupations had higher COVID-19 cumulative incidence. During the uptick of the “Delta” COVID variant (stratified period July 1-Dec 25), counties with more transportation occupations had significantly more COVID-19 cumulative incidence than before.

**Significance:** Understanding the association between occupations and COVID-19 cumulative incidence on an ecological level can provide information for precision public health strategies for prevention and protecting vulnerable workers.

**Impact Statement:** We used data from US Census and COVID-19 data to explore the association between occupations and COVID-19 cumulative incidence and vaccination rate on an ecological level, which can provide information for precision public health strategies for prevention of spread of disease and protecting vulnerable workers.

## Introduction

The Coronavirus Disease 19 (COVID-19) caused by the Severe Acute Respiratory Syndrome coronavirus-2 (SARS-CoV-2) has continued to spread. According to the Centers for Disease Control and Prevention (CDC), the total COVID-19 confirmed case number in the United States has reached 51,574,787 on December 23, 2021 (1). More COVID-19 cases were probably not captured due to limited testing availability (2). It has been postulated that social-economic status, ability to work remotely, and built environment are determinants of the infection rate (3, 4). Thus occupation becomes a risk factor for COVID-19. There is limited evidence on the spread of COVID-19 in different occupations among the general US population. Several research papers have shown that healthcare and other essential workers were more likely to be infected by SARS-CoV-2, but the risk of different non-essential workers in the general US population getting COVID-19 remains unknown (5-7). People in different occupations certainly face different levels of risk of COVID-19, depending on the workplace settings, frequency of interaction with others, ventilation environment, mask-use precautionary measures, and the ability to shelter-in-place when the local government issues such policy (8, 9). Different shelter-in-place regulations in different states or counties may cause the geographic variation in COVID-19 infection, mediated by occupational distribution.

Simultaneously, for the COVID-19 vaccine rollout, some occupations, such as healthcare practitioners and other frontline essential workers, rank high on the list of vaccination priorities (10). Vaccine eligibility has expanded to everyone aged 5 and older in the US. Nevertheless, the difference in COVID-19 vaccine hesitancy has been observed in the general population and specific occupation groups, for example, health care personnel (11-13). Therefore, there can be a disparity in vaccination rates among different occupations. Meanwhile, the disparity in vaccination exists among different counties. As of December 23, 2021, 39% of the counties whose percent of total population with at least one dose is below 50% and only 15% of the counties above 70% (14).

Since July 3, 2021, the Delta variant of SARS-CoV-2 has been estimated to be the predominant lineage in the US (15). The Delta variant is more contagious and it is postulated that the Delta variant might cause more severe illness than previous variants in the unvaccinated (16). According the CDC, from July 3 to December 11, 2021, the proportion of new cases attributed to the Delta variant is predicted to rise from 51.7% to 99% and remains the predominant variant. (15, 17) Therefore, we assumed that the COVID-19 cases were mainly driven by the Delta variant from July 1 to December 25, 2021 and we did a stratified analysis to see the differential effects of occupational variables during the spread of the Delta variant. Upon the emergence of the Omicron variant, retrospective analysis on the Delta variant is still crucial to the comparison between the two variants and the prediction of the future spread of Omicron. The Omicron variant has been detected in most of the states of the US (18). Research has shown that increased risk of SARS-CoV-2 reinfection was associated with the emergence of the Omicron variant (19). Analyses on the occupational determinants of the spread of COVID-19 can be used to inform policy decisions of emerging variants.

Utilizing the geographical variations in COVID-19 case rate and percentage vaccinated, we aim to assess the occupational determinants of COVID-19 infection and vaccination with an ecological analysis of county-level data in the United States accumulated from January 22, 2020 to December 25, 2021. We also evaluated whether our estimates for occupational determinants differed during the spread of the Delta variant from July 1 to December 25, 2021.

## Subjects and Methods

### County-level COVID-19 Cases and Vaccination percentages

We obtained the county-level confirmed cases accumulated from January 22, 2020 up to December 25 2021, from Johns Hopkins Coronavirus Resource Center data repository. The COVID-19 data repository is operated by the Center for Systems Science and Engineering (CSSE) at Johns Hopkins University. To construct a US county-level dataset and display the cumulative COVID-19 confirmed cases on a daily basis, they aggregated data from the Department of Health of states, counties, and cities. We calculated COVID-19 cases per 100,000 using county-level COVID-19 case number and county population from American Community Survey (ACS) 2019 5-year estimate. We collected data for 3194 counties across all 50 states and Puerto Rico. Some counties in Utah and Massachusetts were missing.

We collected the percentage of people who were fully vaccinated as of December 23, 2021 in 3195 counties from the Centers for Disease Control and Prevention (CDC), with missing data in some counties in Massachusetts (Barnstable, Dukes, and Nantucket counties) and in Virginia. Data represents all vaccine partners, including jurisdictional partner clinics, retail pharmacies, long-term care facilities, dialysis centers, Federal Emergency Management Agency and Health Resources and Services Administration partner sites, and federal entity facilities. We used county-level COVID-19 cases per 100,000 and the percentage of people who were fully vaccinated as dependent variables.

### Occupational Data Ascertainment

We collected the percentage of people employed by occupation using the American Community Survey (ACS) 2019 5-year estimate. ACS uses monthly samples to produce annually updated estimates for areas based on the decennial census. ACS questionnaires collected one job from respondents and categorized it into occupation classifications derived from the Standard Occupational Classification (SOC) manual: 2018.

Percentages of people employed by occupation in every county were estimated and we included the 23 broad occupational categories for analysis. We merged health diagnosing and treating practitioners and other technical occupations, health technologists and technicians, and healthcare support occupations into one single category as health-related occupations. Data for 3220 counties were available and all the percentage occupations added up to 100% for every county.

### Covariates

Demographic, socio-economic, environmental, and health-related variables were controlled in regression models. ACS 2019 5-year estimate provided county-level population, population density, age distribution, percentage of females, race distribution, percentage of crowded households, median household income, median housing value, percentage of high school graduates, and percentage of households with an internet subscription. We used data from the PLACES Project launched by the Centers for Disease Control and Prevention (CDC) to control for the prevalence of smoking, chronic obstructive pulmonary disease (COPD), diabetes, obesity, hypertension, and uninsured adults. We also added as a covariate percentage of taking hypertension control medication among those with high blood pressure. The Bureau of Labor Statistics provided county-level unemployment rates. For environmental determinants, we used National Walkability Index from Environmental Protection Agency, Rural-Urban Continuum Codes of the U.S. Department of Agriculture, county-level long term PM2.5 level, the average daily highest temperature in summer and relative humidity in summer from an ecological analysis of air pollution and COVID-19 mortality (20). We excluded samples with missing covariates, and the final numbers of counties in our analysis on COVID-19 cases per 100,000 and percentage vaccinated were 3050 and 3074, respectively.

### Statistical Analysis

Table 1 shows all the data source we used for this paper. We described the distributions and ranges of all variables and used the Pearson correlation to see the association between each occupation percentage and other variables. We fit multivariate linear models for the regression analysis and a total of n=23*2*3=138 models were used in our main analyses. We estimated the effects of every 10% increase in the percent of people employed by occupation by putting every occupational variable into the regression model separately. The outcomes were county-level COVID-19 cumulative incidence and percent of total population fully vaccinated. For every pair of the occupational variable and the outcome, three sets of covariates were added into the regression model stepwisely. Model 1 was adjusted for demographic variables (percentage aged 0-14; percentage aged 15-24; percentage aged 25-34; percentage aged 35-44; percentage aged 45-54; percentage aged 55-64; percentage female; percentage Hispanic; percentage black; percentage Asian; percentage of other races; quintile dummies for county population density; state fixed effects). Model 2 was adjusted for demographic and socio-economic variables (percentage of crowded households; percentage of high school graduates; percentage of uninsured adults; percentage of households with internet subscription; unemployment rates; quintile dummies for median household income; quintile dummies for median housing value). Model 3 was additionally adjusted for health and environmental variables (prevalence of smoking, COPD, diabetes, obesity, and hypertension; percentage of taking hypertension control medication among those with high blood pressure; county-level PM2.5 level; long-term average daily highest temperature in summer and relative humidity in summer; county-level National Walkability Index; 8 dummies for Rural-Urban Continuum Codes).

For the stratified analysis, we selected from July 1 to December 25 as the delta variant predominant period and calculated the COVID-19 cumulative incidence during this period by subtracting the accumulated case number of July 1, 2021 from the accumulated case number of December 25, 2021. For the stratified COVID-19 cumulative incidence, we used the same denominator of county population as above. We also fit the same multivariate linear models and adjusted the same covariates as above. We did all our analyses with SAS OnDemand for Academics.

## Results

We analyzed 3050 counties in the USA in our analysis on COVID-19 cumulative incidence and 3074 counties for COVID-19 vaccination rates. Means for most of these variables were close to their median except population density, median household income, and median housing value (Table 2). Therefore, these three variables were transformed into quintiles to put into the regression model (Table 3). In supplemental Table 1, we calculated the Pearson correlation between each occupation percentage and other variables. The strongest coefficient effect estimate was 0.65, between business and financial operations occupation percentage and median household income. Most of the Pearson coefficients were below 0.5, so it was acceptable for the linear regression model.

Office and administrative support roles, community-based work, production workers, sales occupations, material moving occupations and health professions were related to higher COVID cumulative incidence (p<0.05) (Figure 1). Many professionals such as computer programmers, managers, engineers, scientists, lawyers, agricultural and construction occupations were associated with fewer COVID infections (p<0.05). For example, every 10% increase in health-related occupations were associated with an increase of 1581.1 (95% CI: 1132.1, 2030.1) in COVID-19 cumulative incidence and every 10% increase in computer and mathematical occupations were associated with a decrease of -4539.2 (-5846.7, -3231.7) (Supplemental Table 2). White-collar jobs, food preparation, and protective service occupations were associated with a higher percentage of fully vaccinated populations (p<0.05) (Figure 2 and Supplemental Table 3). Counties with more blue-collar jobs and management occupations had a lower percentage fully vaccinated (p<0.05). Among the occupations with higher COVID-19 infections, community service, material moving, and production workers were associated with lower vaccination rates at the same time. In the stratified analysis covering the Delta variant period, counties with more transportation occupations became negatively associated with COVID-19 cumulative incidence (p<0.05) (Figure 3 and Supplemental Table 4).

**Figure 1.**
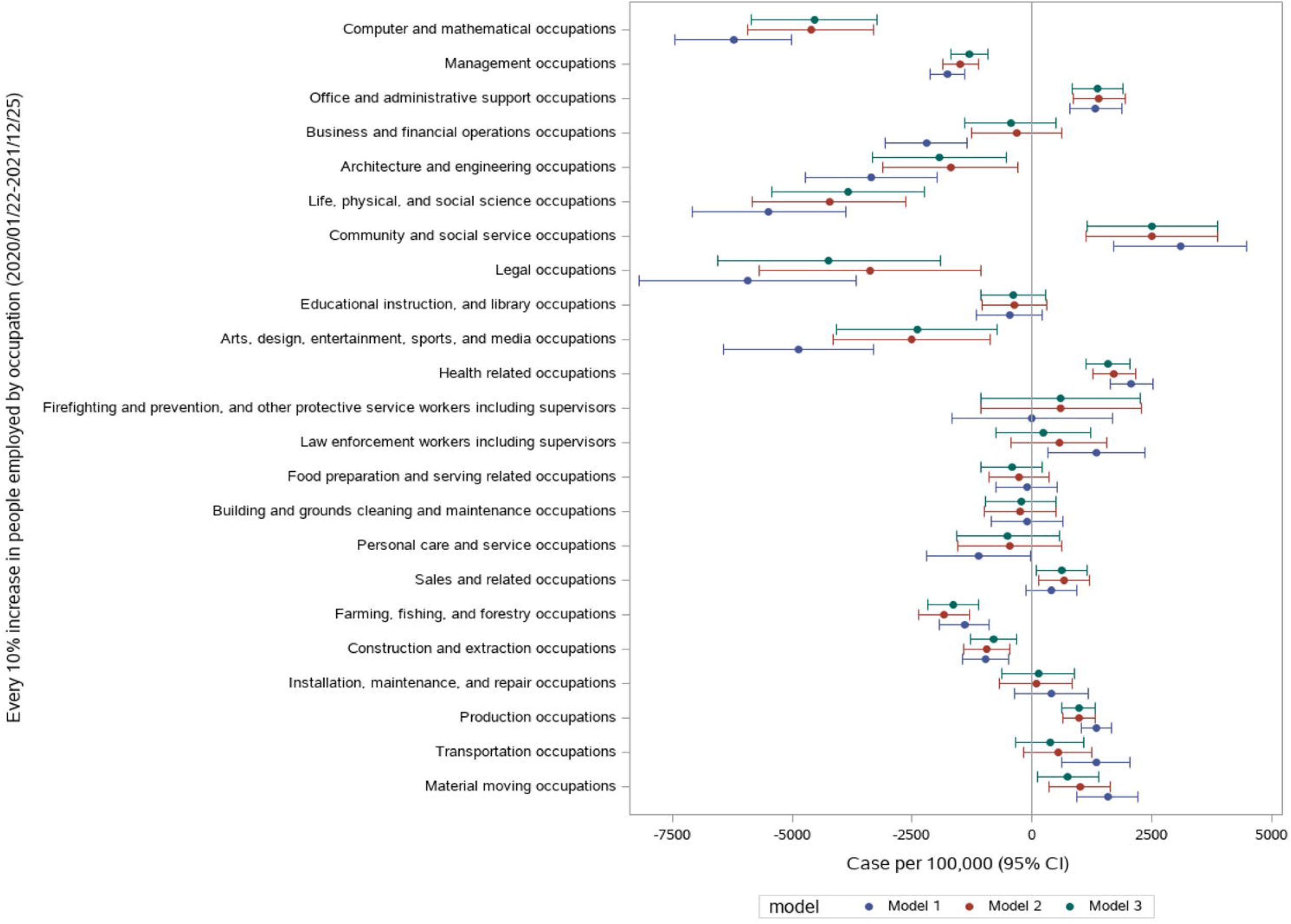
Estimate (95% CI) of association of all occupational variables with COVID-19 cumulative incidence.

**Figure 2.**
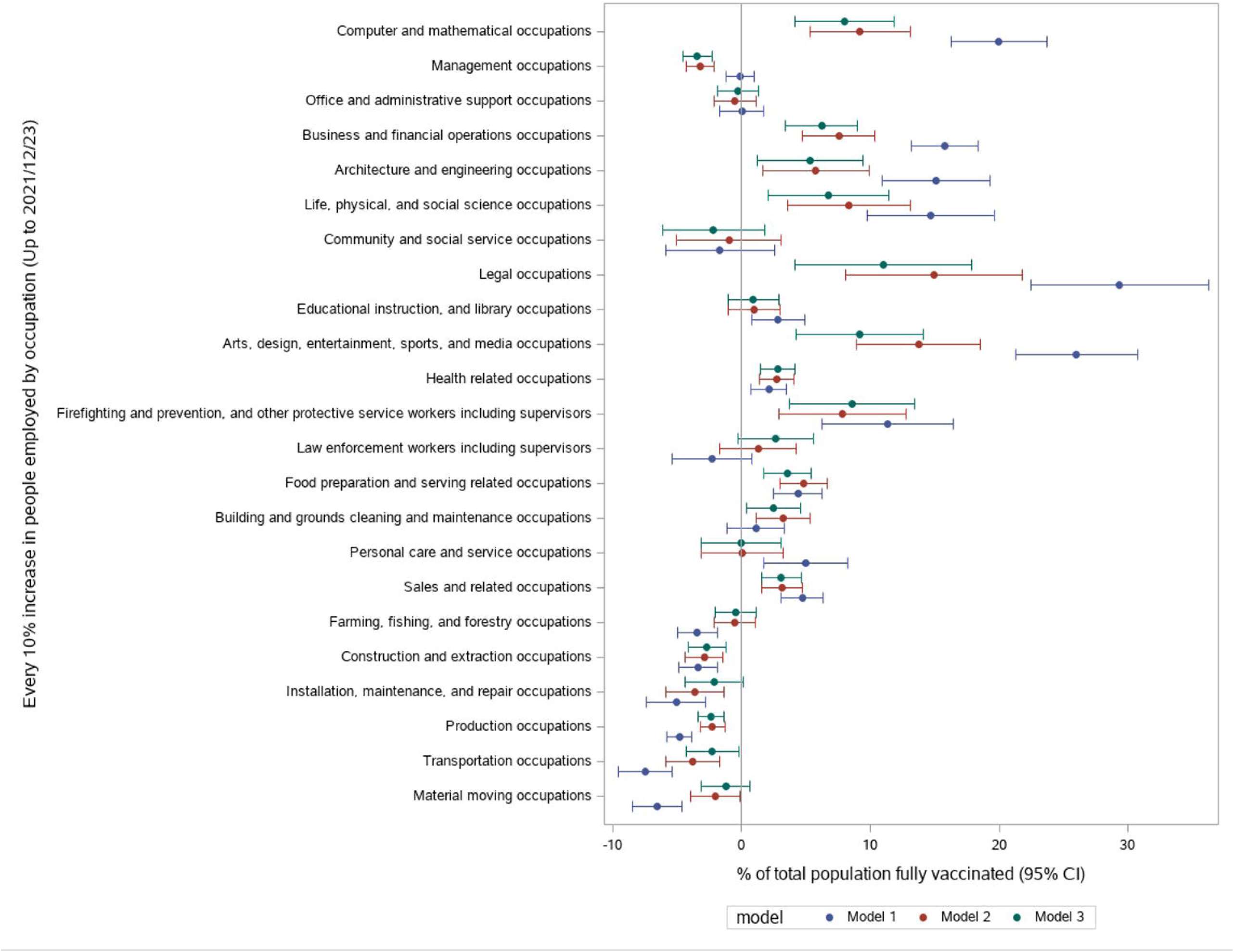
Estimate (95% CI) of association of all occupational variables with percentage fully vaccinated against COVID-19.

**Figure 3.**
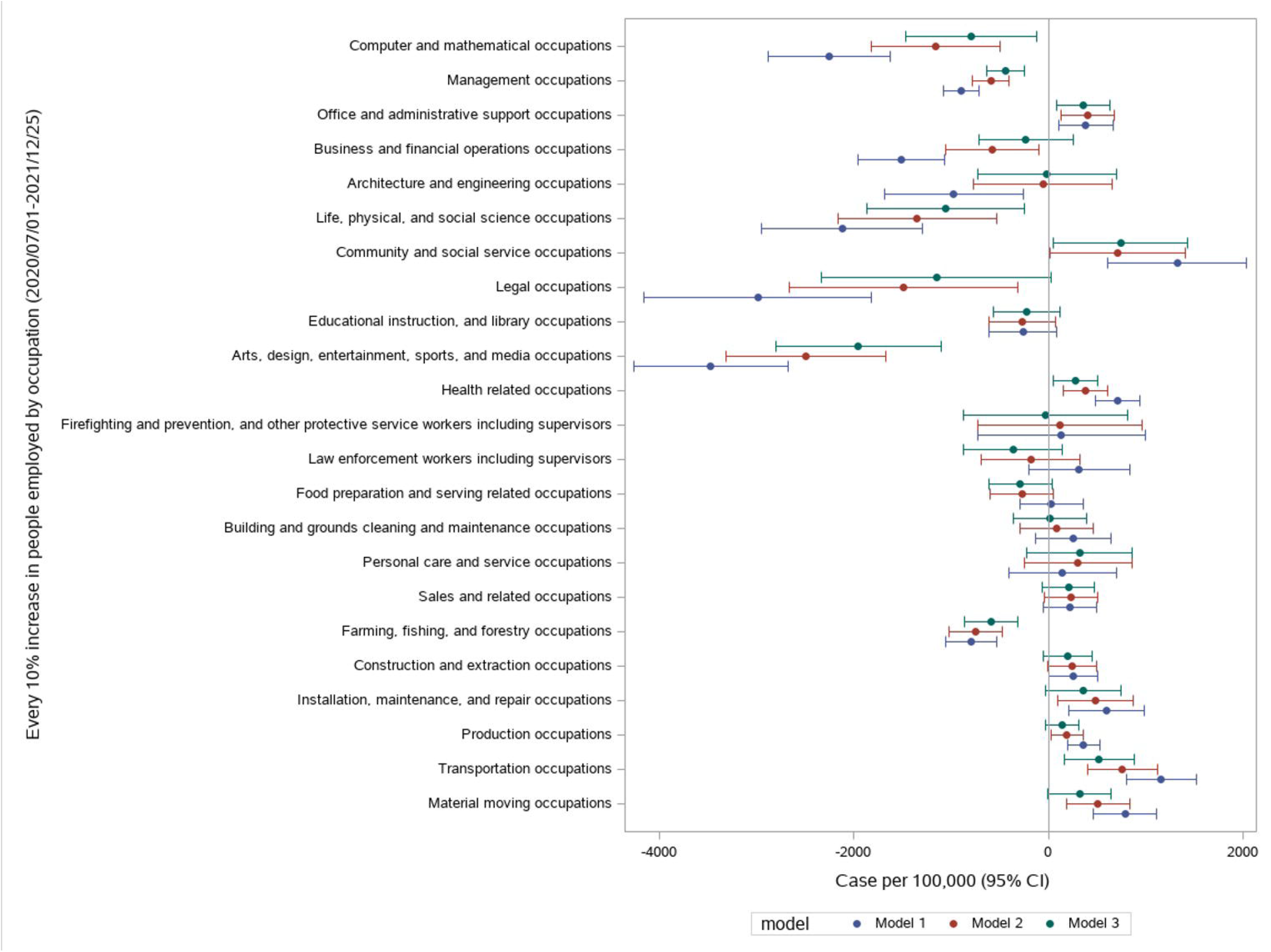
Estimate (95% CI) of association of all occupational variables with COVID-19 cumulative incidence during July 1-Dec 7.

## Discussion

We identified the link between the distribution of occupations and the county COVID-19 cumulative incidence and vaccination rate. Counties with more office supportive roles, community service, health-related jobs, sales, material moving and production occupations had elevated rates of COVID-19 infection. During the stratified period (to analyze the delta variant), counties with more transportation occupations had higher COVID-19 cumulative incidence.

Counties with more professionals such as computer programmers, managers, engineers, scientists, lawyers, and agricultural occupations had a lower rates of COVID-19 infection. Counties with more white-collar jobs, food preparation, and protective service occupations had a higher percentage fully vaccinated. Counties with more blue-collar jobs and management occupations had a lower percentage of those fully vaccinated.

Some research studies have explored the occupational risk of COVID-19. A prospective cohort study conducted in the UK and the USA using data from the COVID Symptom Study smartphone application found that frontline healthcare workers were at increased risk of a positive COVID-19 test, which concurs with our findings (7). An analysis of UK Biobank data found that healthcare workers were associated with a higher risk of being tested for COVID-19 but were not independently associated with the risk of testing positive conditionally on being tested (5). Another analysis of UK Biobank suggested that compared to non-essential workers, healthcare workers, social and education workers, and other essential workers had a higher risk of getting severe COVID-19 (6). Some observational and descriptive studies also found that in Asian healthcare workers, drivers and transport workers, services and sales workers, cleaning and domestic workers and public safety workers had more COVID-19 cases and in Massachusetts healthcare support and transportation and material moving occupations had higher mortality rate (21, 22). Using Occupational Information Network, other researchers estimated and modeled the COVID-19 risk of different occupations, suggesting that apart from health related occupations, protective service occupations, office and administrative support occupations, education occupations, community and social services occupations, and construction and extraction occupations can also be exposed to COVID-19 (8, 9).

The literature in COVID-19 vaccination has also been rapidly growing. Some research has explored the determinants of COVID-19 vaccination hesitancy. A systematic review on global COVID-19 vaccination acceptance found significant geographical and demographic differences in terms of vaccine hesitancy in the general population and the specific subgroups (23). Among health care personnel (HCP), a cross-sectional study concluded that although the majority of HCP were vaccinated, many ancillary workers are still hesitant, which could be risky according to our finding that both health-related and supportive occupations are facing a higher risk of COVID-19 infection (13). Another study on COVID-19 vaccine hesitancy among health care workers found that vaccine hesitancy was highest among black and Hispanic or Latino health care workers (11). Some other researchers explored the optimal allocation of COVID-19 vaccines. One paper determined the age-occupation groups with the top vaccine priority, such as emergency medical technicians and paramedics, nurses, bus drivers, and meat and fish processing workers (24). It is assumed that firefighters at the wildfire incidents were more susceptible to SARS-CoV-2 infection because of the close living and working conditions, limited hygiene supplies, arduous work and environmental exposure to wildfire smoke (25).

Our findings show that apart from health-related occupations, the county-level percent of people employed in other occupations were also associated with an elevated rate of COVID-19 infection. Workers in supportive office roles, community services, sales, material moving and production occupations have a close distance to colleagues or clients and have limited ability to work from home (3, 8). For example, assembly line workers are unlikely to work remotely by now, and they may have to work together in a confined working room. It is also suggested that counties with more community service, material moving and production workers experienced higher level of COVID-19 infection and lower percentage of total population fully vaccinated. Occupational Safety and Health Administration of the US have issued a rule that requires workers at large companies to be vaccinated against COVID-19 or undergo weekly testing, and we suggested that if more workers in occupations with higher risk of COVID-19 infection are fully vaccinated, COVID-19 infection at the workplace settings would be effectively curtailed (26).

Counties with more professionals such as computer programmers, businesspersons, engineers, scientists, and lawyers were often associated with a higher percentage fully vaccinated. Several possible reasons could be able to explain. First, professionals in these occupations may have more access and perhaps a better understanding of COVID-19 information and vaccine effectiveness and side effects. Second, they may have a more flexible schedule that helps book vaccine appointments. They probably have better assess to vaccine uptake as these professions may live in areas with lower commute time to medical resources. Finally, they usually have more income to buffer the potential financial loss due to vaccination, such as delay of work and side effects.

Our study has several strengths. We have included various types of occupations, estimating the effects of 23 occupation variables. We have also explored the association between occupational distribution and vaccination at the ecological level, using data of thousands of counties in the United States. Besides, different kinds of covariates including demographic, socio-economic, environmental, and health-related variables were adjusted in our regression models step by step, serving as control variables and sensitivity analysis. Our analysis of the delta variant impact is unique as no published studies have examined the association of COVID-19 infection caused by the delta variant with occupations.

Limitations also exist in our analysis. First, this is an ecological study which cannot prove causality. Second, underestimation of COVID-19 incidences may be related to occupation classification. Occupations with lower average income may have a higher possibility of not getting tested for COVID-19. Governments and public health agencies in some poorest areas do not have enough resources to conduct testing and vaccinations. These areas may also have more employees in occupations with a higher risk of COVID-19. Third, the COVID-19 pandemic is still ongoing, with the delta variant threatened to cause more cases, especially in counties with low vaccination rates, which may underestimate the county-level accumulated cases. Fourth, some counties with missing vaccination rates possibly have a lower percentage fully vaccinated. Excluding these counties may underestimate the negative effect size of some occupations with less vaccination. We also did not control for physical distancing and other public health interventions, which protects against COVID-19 (27, 28). Counties with more high-risk occupations may also have relaxed public health policies and therefore, physical distancing and other public health interventions become a confounder. While we control for a wide range of confounding covariates, including other measures of socioeconomic status, residual confounding may be a concern. We attempted to control for mobility by including National Walkability Index as an environmental covariate. Besides, county-level covariates may perform poorly in adjusting for confounders since COVID-19 spreads from person to person. Modifiable Areal Unit Problem (MAUP) may also exist in our ecological data. The timing of our data sources were not up-to-date and might be subject to bias caused by rapid changes of variables.

## Conclusion

Our findings indicate that occupational determinants of health exists for COVID-19 infection and vaccine uptake. Our research is informative for targeted public health approach in dealing with the pandemic. The workplace setting plays a part in the spreading of COVID-19. People in different occupations are subject to different levels of COVID-19 infection and social-economic status, ability to work remotely, and the built environment of workplace contribute to the differential occupational risk. Therefore, it is important to promote vaccination in some occupations that are more susceptible to COVID-19 and track the occupation of people getting COVID-19 and vaccination to have a better understanding of the occupational determinants of COVID-19 infection and vaccine uptake.

## Supporting information

Title Page

## Data Availability

All data produced in the present work are gathered from public sources.

## Acknowledgements

The investigators would like to acknowledge the Center for Systems Science and Engineering (CSSE) at Johns Hopkins University and the Centers for Disease Control and Prevention (CDC) for the public data used in this analysis.

## Tables and Figures

Table 1. Data source of all the analysis.

Table 2. Descriptive statistics of all continuous variables.

Table 3. Descriptive statistics of all categorical variables.

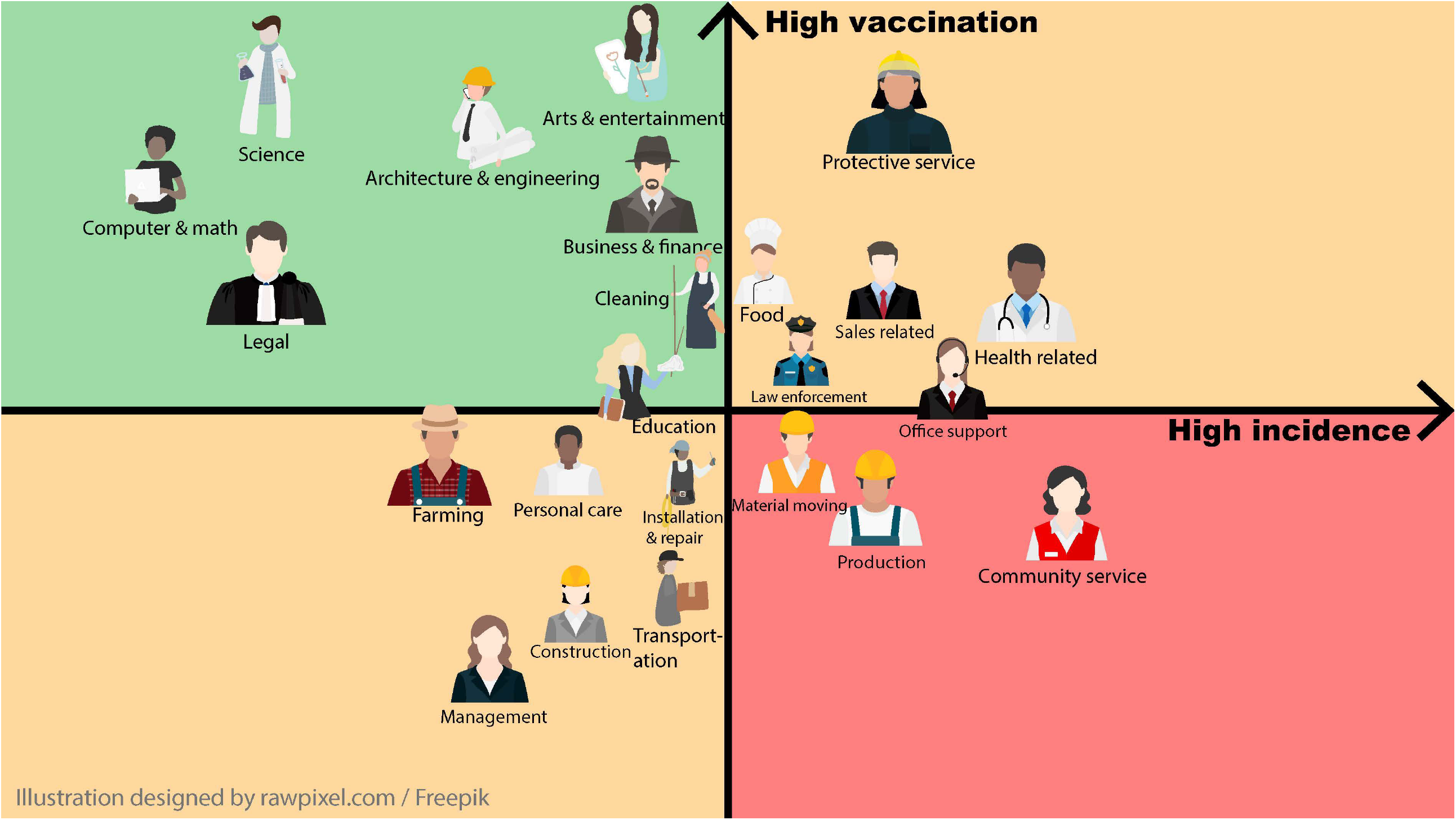

