## Supplementary material for "Occupational determinants of COVID-19 cases and vaccination: an ecological analysis of counties in the United States as of December 2021": Title Page

**Financial disclosure:** No financial disclosures were reported by the authors of this paper.

**Funding:** no financial assistance was received in support of the study.

**Acknowledgements**

The investigators would like to acknowledge the Center for Systems Science and Engineering (CSSE) at Johns Hopkins University and the Centers for Disease Control and Prevention (CDC) for the public data used in this analysis.

**Runing Head Title:** Occupational determinants of COVID-19

**Author Contributions**

**Yucheng Wang:** Data curation, Formal analysis, Investigation, Methodology, Software, Validation, Visualization, Writing – original draft **Dustin T. Duncan:** Investigation, Methodology, Writing – review & editing **John S. Ji:** Conceptualization, Funding acquisition, Investigation, Methodology, Project administration, Resources, Supervision, Validation, Visualization, Writing – review & editing
