## Supplementary material for "Occupational determinants of COVID-19 cases and vaccination: an ecological analysis of counties in the United States as of December 2021": Table 1

Table 1. Data source of all the analysis.

| data source | variables | time period | sample size |
| --- | --- | --- | --- |
| Johns Hopkins Coronavirus Resource Center | County-level COVID-19 cumulative cases | 2020/01/22-2021/12/25 | 3194 |
| Centers for Disease Control and Prevention | Percent of total population fully vaccinated | Up to 2021/12/23 | 3195 |
| American Community Survey 2019 5-year estimate | Percent of people in a certain occupation among all employed | 2014/01/01-2018/12/31 | 3220 |
|  | percentage aged 0-14 |  |  |
|  | percentage aged 15-24 |  |  |
|  | percentage aged 25-34 |  |  |
|  | percentage aged 35-44 |  |  |
|  | percentage aged 45-54 |  |  |
|  | percentage aged 55-64 |  |  |
|  | percentage female |  |  |
|  | percentage Hispanic |  |  |
|  | percentage black |  |  |
|  | percentage Asian |  |  |
|  | percent of other races |  |  |
|  | population |  |  |
|  | percentage of crowded households |  |  |
|  | percentage of high school graduates |  |  |
|  | percentage of households with internet subscription |  |  |
|  | median household income |  |  |
|  | median housing value |  |  |
| PLACES Project launched by CDC | prevalence of smoking | 2021 release | 3121 |
|  | prevalence of chronic obstructive pulmonary disease (COPD) |  |  |
|  | prevalance of diabetes |  |  |
|  | prevalence of obesity |  |  |
|  | prevalence of hypertension |  |  |
|  | percent of uninsured adults |  |  |
|  | percentage of taking hypertension control medication among those with high blood pressure |  |  |
| The Bureau of Labor Statistics | county-level unemployment rate | 2020 | 3141 |
| Environmental Protection Agency | National Walkability Index | 2019 | 3231 |
| Economic Research Service  U.S. Department of Agriculture | Rural-Urban Continuum Codes | 2013 | 3232 |
| Public Available Code and Data to Reproduce Analyses in "Air pollution and COVID-19 mortality in the United States: Strengths and limitations of an ecological regression analysis." | county-level long term PM2.5 level | 2000-2018 | 3097 |
|  | the average daily highest temperature in summer | 2000-2016 | 3108 |
|  | relative humidity in summer | 2000-2016 | 3108 |
