## Supplementary material for "Occupational determinants of COVID-19 cases and vaccination: an ecological analysis of counties in the United States as of December 2021": Table 2

Table 2. Descriptive statistics of all continuous variables.

| **Characteristics** | **N** | **Mean** | **Std Dev** | **Median** | **Minimum** | **Maximum** |
| --- | --- | --- | --- | --- | --- | --- |
| **COVID-19 cumulative incidence 2020/01/22-2021/12/25 (cases per 100,000)** | 3050 | 16961.88 | 3743.33 | 17165.54 | 2811.53 | 59375 |
| **COVID-19 cumulative incidence 2020/07/01-2021/12/25 (cases per 100,000)** | 3050 | 6638.84 | 1911.3 | 6609.63 | 803.21 | 16553.03 |
| **COVID-19 percentage fully vaccinated (%)** | 3074 | 46.85 | 12.72 | 46.50 | 0.00 | 95.00 |
| **Occupation variables (%)** |  |  |  |  |  |  |
| **Computer and mathematical occupations** | 3074 | 1.44 | 1.3 | 1.13 | 0 | 13.16 |
| **Management occupations** | 3074 | 10.02 | 3.9 | 9.2 | 0 | 45.54 |
| **Office and administrative support occupations** | 3074 | 11 | 1.89 | 11.02 | 2.33 | 22.26 |
| **Business and financial operations occupations** | 3074 | 3.4 | 1.59 | 3.18 | 0 | 14.55 |
| **Architecture and engineering occupations** | 3074 | 1.28 | 0.88 | 1.17 | 0 | 11.75 |
| **Life, physical, and social science occupations** | 3074 | 0.72 | 0.68 | 0.6 | 0 | 19.09 |
| **Community and social service occupations** | 3074 | 1.75 | 0.78 | 1.69 | 0 | 8.01 |
| **Legal occupations** | 3074 | 0.59 | 0.52 | 0.5 | 0 | 7.11 |
| **Educational instruction, and library occupations** | 3074 | 6.05 | 1.78 | 5.82 | 0 | 18.18 |
| **Arts, design, entertainment, sports, and media occupations** | 3074 | 1.15 | 0.81 | 1.03 | 0 | 8.19 |
| **Health related occupations** | 3074 | 9.41 | 2.47 | 9.37 | 0 | 21.9 |
| **Firefighting and prevention, and other protective service workers including supervisors** | 3074 | 0.95 | 0.67 | 0.88 | 0 | 9.78 |
| **Law enforcement workers including supervisors** | 3074 | 1.37 | 1.2 | 1.04 | 0 | 17.97 |
| **Food preparation and serving related occupations** | 3074 | 5.44 | 1.79 | 5.32 | 0 | 20.07 |
| **Building and grounds cleaning and maintenance occupations** | 3074 | 4.16 | 1.51 | 3.94 | 0 | 24.76 |
| **Personal care and service occupations** | 3074 | 2.43 | 0.98 | 2.42 | 0 | 14.06 |
| **Sales and related occupations** | 3074 | 9.18 | 2.09 | 9.22 | 0.54 | 22.09 |
| **Farming, fishing, and forestry occupations** | 3074 | 2.05 | 2.62 | 1.16 | 0 | 38.86 |
| **Construction and extraction occupations** | 3074 | 6.45 | 2.48 | 6 | 0.32 | 24.39 |
| **Installation, maintenance, and repair occupations** | 3074 | 4.12 | 1.48 | 4.04 | 0 | 23.2 |
| **Production occupations** | 3074 | 8.18 | 4.22 | 7.49 | 0 | 30.03 |
| **Transportation occupations** | 3074 | 4.55 | 1.59 | 4.36 | 0 | 17.59 |
| **Material moving occupations** | 3074 | 4.28 | 1.84 | 4.1 | 0 | 24.93 |
| **Demographic** |  |  |  |  |  |  |
| **Age (%)** |  |  |  |  |  |  |
| **0-14** | 3074 | 18.34 | 3.01 | 18.3 | 6 | 35.9 |
| **15-24** | 3074 | 12.57 | 3.39 | 12 | 1.3 | 44.8 |
| **25-34** | 3074 | 11.81 | 2.24 | 11.6 | 3.4 | 27.2 |
| **35-44** | 3074 | 11.58 | 1.52 | 11.6 | 4 | 19.2 |
| **45-54** | 3074 | 12.61 | 1.47 | 12.7 | 5.3 | 19.5 |
| **55-64** | 3074 | 14.23 | 2.15 | 14.2 | 5.2 | 29.8 |
| **female (%)** | 3074 | 49.94 | 2.31 | 50.4 | 27.3 | 56.1 |
| **Hispanic (%)** | 3074 | 9.41 | 13.95 | 4.2 | 0 | 99.2 |
| **black (%)** | 3074 | 8.95 | 14.47 | 2.2 | 0 | 87.2 |
| **Asian (%)** | 3074 | 1.24 | 2.27 | 0.6 | 0 | 36.3 |
| **Other races (%)** | 3074 | 1.72 | 6.36 | 0.5 | 0 | 87.4 |
| **Population density (person per sq mile)** | 3074 | 253.82 | 1790.83 | 44.64 | 0.22 | 72052.96 |
| **Socio-economic** |  |  |  |  |  |  |
| **Median household income ($/year)** | 3074 | 53137.43 | 13826.34 | 51616.5 | 21504 | 142299 |
| **Median housing value ($)** | 3074 | 151600.85 | 93326.05 | 126450 | 24400 | 1097800 |
| **High school graduate and above (%)** | 3074 | 86.9 | 6.27 | 88.26 | 26.44 | 98.88 |
| **Unemployment (%)** | 3074 | 6.68 | 2.21 | 6.5 | 1.7 | 22.5 |
| **Internet subscription (%)** | 3074 | 75.92 | 8.74 | 77.1 | 35.2 | 96.3 |
| **Environment** |  |  |  |  |  |  |
| **Average PM2.5 from 2000-2016 (µg/m3)** | 3074 | 8.38 | 2.52 | 8.77 | 2.06 | 15.79 |
| **Summer temperature from 2000-2016 (℃)** | 3074 | 29.98 | 3.18 | 30.17 | 17.31 | 40.72 |
| **Summer relative humidity from 2000-2016 (%)** | 3074 | 88.95 | 9.73 | 91.33 | 31.64 | 99.78 |
| **County-level Walkability Index (0-20)** | 3074 | 6.46 | 1.87 | 6.08 | 2.86 | 16 |
| **Crowded household (%)** | 3074 | 2.31 | 1.79 | 1.86 | 0 | 15.41 |
| **Health related** |  |  |  |  |  |  |
| **Smoking (%)** | 3074 | 20.38 | 4.14 | 20.1 | 6.5 | 38.2 |
| **COPD (%)** | 3074 | 7.41 | 1.83 | 7.2 | 3.2 | 15.5 |
| **Diabetes (%)** | 3074 | 10.79 | 2.33 | 10.4 | 5.5 | 21 |
| **Obesity (%)** | 3074 | 35.8 | 4.25 | 36.2 | 16.4 | 51 |
| **Hypertension (%)** | 3074 | 32.71 | 4.87 | 31.7 | 21.4 | 53 |
| **Taking hypertension control medication among those with hypertension (%)** | 3074 | 57.82 | 3.84 | 57.8 | 46.2 | 71.9 |
| **Uninsured adult (%)** | 3074 | 18.41 | 6.87 | 16.7 | 7.9 | 56.6 |
