## Supplementary material for "Occupational determinants of COVID-19 cases and vaccination: an ecological analysis of counties in the United States as of December 2021": Table 3

Table 3. Descriptive statistics of all categorical variables.

| **Characteristics** | Cases per 100,000 | | Percentage fully vaccinated (%) | |
| --- | --- | --- | --- | --- |
|  | **N (%)** | **Mean** | **N (%)** | **Mean** |
| **Urban Rural Code** |  |  |  |  |
| **1: Counties in metro areas of 1 million population or more** | 413 (13.54) | 15665.22 | 413 (13.44) | 53.17 |
| **2: Counties in metro areas of 250,000 to 1 million population** | 368 (12.07) | 16485.95 | 372 (12.1) | 51.12 |
| **3: Counties in metro areas of fewer than 250,000 population** | 348 (11.41) | 17041.92 | 350 (11.39) | 47.61 |
| **4: Urban population of 20,000 or more, adjacent to a metro area** | 213 (6.98) | 17220.14 | 214 (6.96) | 48.32 |
| **5: Urban population of 20,000 or more, not adjacent to a metro area** | 89 (2.92) | 18133.27 | 89 (2.9) | 49.03 |
| **6: Urban population of 2,500 to 19,999, adjacent to a metro area** | 585 (19.18) | 17650.18 | 587 (19.1) | 44.41 |
| **7: Urban population of 2,500 to 19,999, not adjacent to a metro area** | 413 (13.54) | 17884.62 | 423 (13.76) | 45.05 |
| **8: Completely rural or less than 2,500 urban population, adjacent to a metro area** | 218 (7.15) | 16299.46 | 219 (7.12) | 41.99 |
| **9: Completely rural or less than 2,500 urban population, not adjacent to a metro area** | 403 (13.21) | 16674.55 | 407 (13.24) | 42.6 |
| **Population density quintile (Persons per square mile)** |  |  |  |  |
| **Q1 (<12.36)** | 599 (19.64) | 16682.58 | 614 (19.97) | 43.72 |
| **Q2 (12.36-31.7)** | 611 (20.03) | 17322.41 | 615 (20.01) | 44.6 |
| **Q3 (31.7-62.4)** | 615 (20.16) | 17608.04 | 615 (20.01) | 44.94 |
| **Q4 (62.4-155.3)** | 613 (20.1) | 17172 | 615 (20.01) | 46.28 |
| **Q5 (>155.3)** | 612 (20.07) | 16015.5 | 615 (20.01) | 54.69 |
| **Household income quintile ($)** |  |  |  |  |
| **Q1 (<42341)** | 615 (20.16) | 17594.37 | 615 (20.01) | 42.19 |
| **Q2 (42345-48838)** | 614 (20.13) | 17364.33 | 614 (19.97) | 42.74 |
| **Q3 (48856-54089)** | 613 (20.1) | 17143.95 | 615 (20.01) | 46.16 |
| **Q4 (54090-61633)** | 607 (19.9) | 17081.95 | 615 (20.01) | 49.51 |
| **Q5 (>61655)** | 601 (19.7) | 15610.74 | 615 (20.01) | 53.63 |
| **Housing value quintile ($)** |  |  |  |  |
| **Q1 (<91400)** | 614 (20.13) | 17991.11 | 614 (19.97) | 41.78 |
| **Q2 (91500-114100)** | 612 (20.07) | 17721.47 | 615 (20.01) | 43.66 |
| **Q3 (114300-143500)** | 611 (20.03) | 17409.44 | 615 (20.01) | 44.81 |
| **Q4 (143600-190300)** | 607 (19.9) | 17133.4 | 615 (20.01) | 48.81 |
| **Q5 (>190400)** | 606 (19.87) | 14502.91 | 615 (20.01) | 55.17 |
